# Post-exposure Prophylaxis or Preemptive Therapy for SARS-Coronavirus-2: Study Protocol for a Pragmatic Randomized Controlled Trial

**DOI:** 10.1101/2020.05.01.20087999

**Authors:** Sylvain A Lother, Mahsa Abassi, Alyssa Agostinis, Ananta S Bangdiwala, Matthew P Cheng, Glen Drobot, Nicole Engen, Kathy H Hullsiek, Lauren E Kelly, Todd C Lee, Sarah M Lofgren, Lauren J MacKenzie, Nicole Marten, Emily G McDonald, Elizabeth C Okafor, Katelyn A Pastick, Matthew F Pullen, Radha Rajasingham, Ilan Schwartz, Caleb P Skipper, Alexis F Turgeon, Ryan Zarychanski, David R Boulware

## Abstract

**Background:** The severe acute respiratory syndrome coronavirus 2 (SARS-CoV-2) emerged in December 2019 causing the coronavirus disease 2019 (COVID-19) pandemic. Currently, there are a lack of evidence-based therapies to prevent COVID-19 following exposure, or to prevent worsening of symptoms following confirmed infection. We describe the design of a clinical trial of hydroxychloroquine for post-exposure prophylaxis and pre-emptive therapy for COVID-19.

**Methods:** We will conduct two nested multicenter international double-blind randomized placebo-controlled clinical trials of hydroxychloroquine for: 1) post-exposure prophylaxis (PEP) of asymptomatic household contacts or healthcare workers exposed to COVID-19 within the past four days, and 2) pre-emptive therapy (PET) for symptomatic outpatients with COVID-19 with a total symptom duration of less than 4 days. We will recruit 1500 patients for each the PEP and PET trials. Participants will be randomized 1:1 to receive 5 days of hydroxychloroquine or placebo. The primary PEP trial outcome will be the incidence of symptomatic COVID-19 disease. The primary PET trial outcome will be an ordinal scale of disease severity (not hospitalized; hospitalized without intensive care, hospitalization with intensive care, or death). Participant screening, informed consent, and follow up will be exclusively internet-based with appropriate regulatory and research ethics board approvals in Canada and the United States.

**Discussion:** These complementary randomized control trials are innovatively designed and adequately powered to rapidly answer urgent questions regarding the effectiveness of hydroxychloroquine to reduce transmission and disease severity of COVID-19 during a pandemic. In-person participant follow-up will not be conducted in order to facilitate social distancing strategies and reduce risks of exposure to study personnel. Innovative trial approaches are needed to urgently assess therapeutic options to mitigate the global impact of this pandemic.

**Trials Registration:** clinicaltrials.gov (NCT04308668); 16 March 2020.

## Introduction

Coronavirus disease 2019 (COVID-19) is a novel infection caused by Severe Acute Respiratory Syndrome-related Coronavirus-2 (SARS-CoV-2).^1^ The virus was first recognized in Wuhan, China in December 2019 and quickly spread globally, leading to a pandemic.^2^ Currently, the standard of care in most jurisdictions for those with mild to moderate COVID-19 includes self-observation and self-quarantine for 7-14 days.^3,4^ While some small studies^5–8^ with methodologic limitations have suggested certain therapies may have promise, at the time of this publication no proven effective therapies exist. Most major national and international guidelines have yet to incorporate pharmacologic therapies for COVID-19 due to a lack of high grade evidence.^9,10^ An effective post-exposure prophylaxis or pre-emptive therapeutic intervention would provide significant benefit to individuals, as well as to public health, given the high rate of SARS-CoV-2 transmission and overburdened healthcare systems.

The onset of symptoms from COVID-19 generally develops an average of 4-5 days after exposure to SARS-CoV-2.^11^ The incubation period ranges between 1-14 days.^12^ The majority (80%) of people with COVID-19 exhibit mild symptoms and recover spontaneously.^13^ However, 14% of patients develop hypoxemia requiring hospitalization, admission to the intensive care unit (ICU), and/or invasive ventilation.^14^ Of those who are hospitalized in North America, approximately 30% of individuals will develop respiratory failure, shock, and/or multiple organ system dysfunction requiring an admission to a critical care unit.^15^

SARS-CoV-2 has demonstrated rapid spread through the population with a basic reproductive value (R_0_) value of 2 to 5.7.^16,17^ The secondary attack rate among household contacts has been estimated at 10% (95% CI, 2 to 31%),^18^ but estimates are largely based on monitoring of travel-associated COVID-19 cases. In the context of community transmission, the secondary attack rate may be much higher (35%; 95% CI, 27 to 44%).^19^ In the absence of appropriate distancing strategies, secondary attack rates may be significantly higher. Since up to 14% of patients with secondary infections will require hospitalization and are at risk of poor outcomes, including death, a protective therapeutic strategy is urgently needed following exposure to COVID-19.

While few pharmacologic agents have shown *in vitro* suppression of viral replication against coronaviruses^20^, one agent, chloroquine, has been shown to be effective at reducing laboratory established infection and spread to adjacent cells from SARS-CoV.^21^ More recently, the related drug hydroxychloroquine was found to prevent SARS-CoV-2 viral replication before and after infection of Vero cell lines.^22^ The exact anti-viral mechanism of hydroxychloroquine against SARS-CoV-2 is incompletely understood, but may relate to impaired angiotensin converting enzyme-2 (ACE2) receptor binding and resultant cell entry, as well as the suppression of cytokine storm.^23^

Recent news outlet reports of chloroquine poisoning have raised concerns regarding the safety of hydroxychloroquine, but scientific details are lacking. A recent trial of hospitalized COVID-19 patients in Brazil was stopped early (n=81) due to safety concerns. Patients receiving high dose chloroquine (12 grams over 10 days) had prolonged QTc (> 500 msec) in 25%, including 2 deaths secondary to ventricular tachycardia.^24^ This dose was orders of magnitude higher than for malaria prophylaxis (500 mg weekly) or malaria treatment (2.5 grams total), each of which have well-established safety profiles.^25^ Hydroxychloroquine is much less toxic that hydroxychloroquine,^26^ and has decades of established safety data in the treatment of malaria and other autoimmune conditions.^27^ Preliminary data in COVID-19 trials have demonstrated increased GI upset but no significant differences in serious adverse events.^28^

Early translational and clinical studies have demonstrated that hydroxychloroquine might be an effective pharmacologic treatment to prevent viral establishment if used early after exposure or established infection.^5–8,29^ Delaying treatment to the time of hospitalization, which occurs on average 7-8 days from symptom onset,^30,31^ may be associated with a lack of benefit due to established viral infection.^28,32^ However, the current evidence base is limited due to small, underpowered sample sizes and large observational studies without proper controls.^20^ Global political forces continue to emphasize a not-yet-proven role for hydroxychloroquine in the paradigm of COVID-19 treatment. Urgent clinical trials are therefore needed to establish whether hydroxychloroquine may be an effective therapy for COVID-19.

### Trial Objectives

To determine if hydroxychloroquine as compared to placebo is effective at: 1) preventing COVID-19 disease after an exposure to a known case (i.e., post-exposure prophylaxis), and 2) preventing severe complications of the disease if started early in the course of illness (i.e., preemptive therapy).

### Trial design

This trial is a double-blind placebo-controlled multicenter international randomized-controlled trial designed to evaluate the superiority of hydroxychloroquine versus placebo. Participants will be enrolled in two nested trials that will be randomized separately and have separate endpoints but use the same workflow, study team, documents and infrastructure. The post-exposure prophylaxis (PEP) trial will include asymptomatic healthcare workers or household contacts exposed to COVID-19 positive cases within 4 days. The pre-emptive therapy (PET) trial will include outpatients with COVID-19 and symptoms for 4 or fewer days duration. Each trial is planned to recruit a total of 1500 patients. Within each trial, participants will be randomized 1:1 in parallel groups to receive hydroxychloroquine or placebo. This clinical trial was registered at Clinical Trials.gov (NCT<u>04308668) on 16 March 2020</u> and was approved by the Food and Drug Administration (IND 148257) and Health Canada. Research ethics board (REB) approval was obtained at the University of Minnesota (16 March 2020), McGill University Health Centre Research Institute (1 April 2020), University of Alberta (6 April 2020), and University of Manitoba (25 March 2020), prior to recruitment of the first participant.

## Methods and Analysis

### Trial setting

Participants will be non-hospitalized adults recruited from the community via advertisements and using traditional methods and social media (Twitter, Instagram, Facebook). Recruitment will take place throughout the United States, and, at least initially, within three provinces in Canada (Quebec, Manitoba, Alberta). Approval is being sought in additional provinces. In order to limit the spread of disease, no in-person visits with study personnel will be performed. During recruitment and follow-up, most participants will be at home in self isolation or quarantine; screening, informed consent, randomization and data collection will occur online through the Research Electronic Data Capture (REDCap), a secure web-based platform. Study medications will be delivered directly to the participants’ place of residence via courier services that respect safety protocols for home deliveries to patients with COVID-19. Participants can contact the local study teams in their region via email or phone for questions or concerns with timely responses (same day during business hours and the following day after 9pm). Websites have been created that will provide live enrollment updates, responses to frequently asked questions, and trial information for participants (www.covidpep.umn.edu and www.covid-19research.ca).

### Inclusion criteria

- Adults (≥ 18 years old)
- Able to read and comprehend in French or English
- Access to the internet with valid email address for enrollment and follow-up surveys

Post-exposure prophylaxis (PEP)
1. At-risk exposure to a COVID-19 positive case within the past 4 days
2. Household contact *<u>or</u>* healthcare worker

Pre-emptive therapy (PET)
1. Non-hospitalized community-dwelling symptomatic (fever, cough, or shortness breath) COVID-19 disease within 4 days of symptom onset
2. COVID-19 disease confirmed by PCR *or* for health care workers, by compatible symptoms with exposure to a known PCR positive case (exposure occurring within the past 14 days)

Enrollment for the PEP trial will be limited to asymptomatic individuals exposed to a COVID-19 confirmed case within ≤ 4 days, as the mean incubation period for SARS-CoV-2 is 5 days.^12^ Household contacts are defined as residing in the same domicile. At risk exposures in the healthcare setting are defined as greater than 10 minutes spent within 2 meters of a COVID-19 confirmed case, with insufficient use of personal protective equipment per institutional guidelines. Healthcare workers are self-identified physicians, nurses, respiratory therapists, or other workers providing direct patient contact. This will select a high-risk group for developing clinical infection but within the time period that the intervention could prevent or ameliorate disease. All efforts will be made to deliver study medicines prior to the 4^th^ day following exposure; however, delays may lead to symptom onset after randomization but prior to starting the intervention. These events will be tracked by online surveys administered upon receipt of the intervention. Due to community testing limitations for COVID-19, participants will be included in the PET group as long as symptoms are compatible with COVID-19 after exposure to a person with confirmed COVID-19.

### Exclusion criteria

1. Known allergy to chloroquine or hydroxychloroquine
2. Current use of chloroquine or hydroxychloroquine for any reason
3. Currently hospitalized
4. Known chronic kidney disease, stage 4-5, or dialysis^33^
5. Active malignancy on systemic chemotherapy
6. Known porphyria
7. Prior retinal eye disease
8. Known glucose-6-phosphate dehydrogenase (G6PD) deficiency
9. Weight < 40 kg
10. Known history of ventricular arrythmia, prolonged QTc interval (personal or family history), structural or ischemic heart disease, or any known episode of sudden cardiac death
11. Severe diarrhea and/or vomiting at screening that may interfere with drug absorption
12. Significant hepatic impairment defined as known cirrhosis with a history of hepatic encephalopathy or ascites
13. Known pregnancy or breastfeeding
14. Current use of any of the following medications with known drug-drug interactions: amiodarone, amitriptyline, artemether, azithromycin, ciprofloxacin, citalopram, clarithromycin, dapsone, desipramine, digoxin, dofetilide, doxepin, droperidol, erythromycin, escitalopram, flecainide, fluoxetine, haloperidol, imipramine, itraconazole, ketoconazole, levofloxacin, lithium, lumefantrine, mefloquine, methadone, moxifloxacin, procainamide, propafenone, quetiapine, sertraline, sotalol, sumatriptan, thioridazine, venlafaxine, bupropion, ziprasidone, zolmitriptan
15. Current use of any anti-epileptic

Exclusion criteria 11-14 are specific only to participants in Canada at the request of Health Canada. Exclusion criteria 15 is specific only to participants in the US. In the US, the FDA has approved the inclusion of pregnant and breastfeeding women citing hydroxychloroquine safety data in pregnant patients with autoimmune conditions including systemic lupus erythematosus and malaria.^34^

### Interventions

Participants will be randomized 1:1 to receive hydroxychloroquine or placebo.

#### Intervention arm

Hydroxychloroquine sulfate will be administered in 200 mg tablets for a total dose of 3,800 mg (19 tablets) over 5 consecutive days. Dosing regimens will consist of a loading dose with subsequent maintenance dose as follows:

1. Day 1 (upon receipt of study medicine) - hydroxychloroquine 800 mg orally once (4 tablets), followed 6 to 8 hours later by 600 mg orally once (3 tablets);
2. Days 2 to 5 -hydroxychloroquine 600 mg orally once daily (3 tablets per day).

#### Control arm

Placebo tablets will be administered using the same dosing schedule and number of tablets as the interventional arm.

The rational for the study dose is adapted from a modified malaria dosing schedule for hydroxychloroquine.^35^ Considerations were made for maximal viral inhibition while balancing potential toxicity and drug supply with anticipated shortages. A loading dose was used, similar to malaria treatment strategy to achieve anti-viral activity as quickly as possible. Monte Carlo simulations were conducted with the proposed dose achieving steady-state serum concentrations above the EC50,^22^ where 50% of viral inhibition would occur.

The most common anticipated side effect from hydroxychloroquine is gastrointestinal upset.^27^ Participants will be instructed to spread out doses throughout the day (e.g., 2 tabs with breakfast, 1 tab with lunch) to increase tolerability if required. Participants may voluntarily stop taking the study medicine at any time. Adherence will be collected by self-report. Chronic medications will be taken as usual, but these must be taken 4 hours before or after taking the study medicine. No other concomitant COVID-19 therapy or other trial medicines are permitted for the duration of the trial. In the event of hospitalization, unblinding of group assignment can occur at the request of the treating physician, and participants may receive other medications or therapies, including experimental treatments from other trials.

### Outcomes

Primary outcome for PEP:

1. Incidence of proven COVID-19 disease onset by 14 days.

In the absence of SARS-CoV-2 testing, COVID-19 diagnosis will be made using clinical criteria and labelled as a possible case. For the “confirmed cases” outcome, there may be limitations related to access to testing. We will use an *a priori* clinical definition using the symptoms most commonly reported in PCR proven patients from the World Health Organization (WHO).^36^

Primary outcome for PET:

1. Ordinal scale for clinical severity, defined by WHO COVID-19 therapeutic trial synopsis^37^
  1 = Ambulatory
  2 = Hospitalized with mild disease
  3 = Hospitalized with severe disease
  4 = Death (all cause)

Secondary outcomes for PEP and PET:

1. Incidence of hospitalization for COVID-19;
2. Incidence of death from COVID-19;
3. Incidence of confirmed SARS-CoV-2 detection;
4. Incidence of possible COVID-19 symptoms;
5. Incidence of all-cause study medicine discontinuation;
6. Severity of symptoms at Day 5 and 14 by visual analogue scale;

Analysis for the primary and secondary outcomes listed will be performed separately for both trials. Assessment of outcome measures will be through self-report via online questionnaires. Wherever possible, COVID-19 disease status will be verified from public health records, medical records, or death certificates. Patients with positive molecular testing (i.e., RT-PCR of nasopharyngeal swab) for SARS-CoV-2 will be considered as confirmed cases.^38^ Molecular testing will be performed as per local institution standard procedures.

For patients with COVID-19 disease who require hospitalization, outcome data will be collected up to day 90. Participants will complete additional online surveys throughout hospitalization to assess for adverse events related to the study medicine and their COVID-19 related outcomes. All participants will complete virtual assessments on days 1, 5, and 14 (**Figure 1**). If symptomatic disease develops, participants will be required to complete additional virtual assessments on day 3 and 10. Participants who are pregnant or become pregnant while participating in the trial will be followed to the post-partum period to assess maternal and fetal outcomes. In the event that a participant does not respond to the day 14 survey, we will contact them by telephone, by registered mail, and if they are still not available, all participants have given a 3^rd^ party telephone contact to verify vital status.

**Figure 1:**
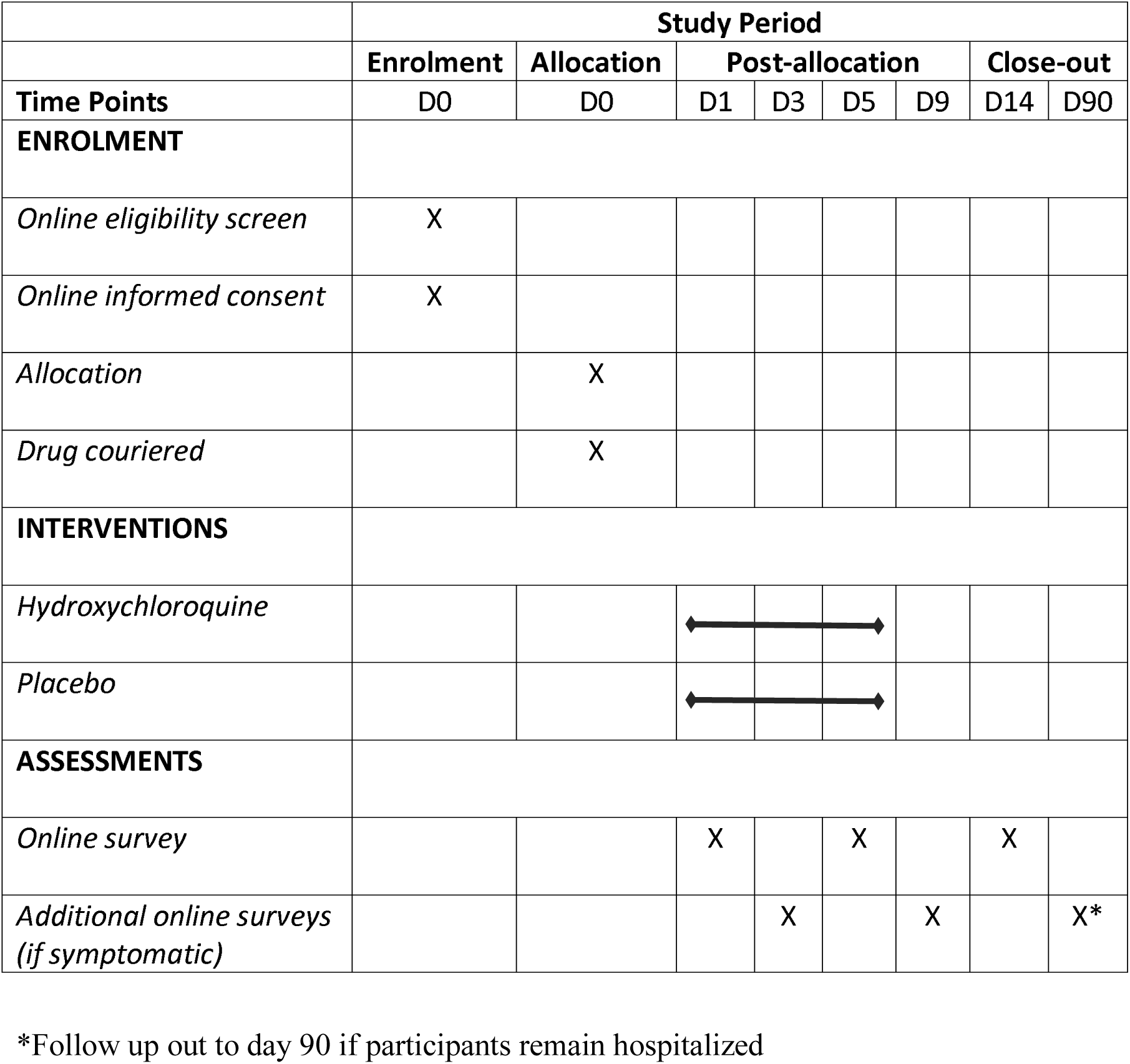
Participant schedule of enrolment, interventions, and assessments

### Recruitment and consent

Recruitment and follow up will be exclusively internet-based. Prospective participants will visit the trial website, read the provided study information, and can click the link to enroll. The trial website will be advertised through communication strategies and media channels, and through internet-based advertisements and social media. In all regions, COVID-19 positive cases are reported to public health where positive patients are notified, and contact tracing is initiated. In some jurisdictions, public health officials will provide information on accessing the trial website. Health care workers will be notified of the trial website by email communications and through the offices of occupational health and promotion by senior hospital administration. Online screening questionnaires will assess interested participants for eligibility criteria. Eligible participants will provide informed consent by submitting an online form, linked through the trial website. Contact information will be provided for participants to ask questions by email and/or phone.

### Randomization sequence generation, allocation concealment, and blinding

Participants will be randomized via computer-generated permuted block randomization and stratified into the PEP trial (for asymptomatic participants) or the PET trial (for symptomatic participants). The randomization sequence will be pre-specified. Randomization will be recorded on an electronic log which will only be accessed by the research pharmacy, those packaging study medication and the unblinded statisticians. Study investigators, trial participants, care providers, outcome assessors, and data analysts will be blinded to the allocation sequence. Once study medication is prepared by the onsite pharmacies, packages will be shipped directly to participants via courier.

Participants will be provided masked study medicine and will not be notified whether they are taking hydroxychloroquine or placebo. Instructions for taking the medicine are identical between the groups. Due to a variety of sources of hydroxychloroquine being used in different jurisdictions, the placebo will not be identically matched to the hydroxychloroquine tablet; however, both placebo and hydroxychloroquine will have similar rounded oval shapes, color, and indistinct markings making unblinding of the trial arm unlikely. Patients will be asked if they believe they received the intervention or control upon trial closure. In the event of a medical emergency, an on-call investigator will be available for code breaking if required. Any code breaking event will be reported to the research pharmacy and principal investigator.

### Data collection methods and data management

All data collection, including baseline demographics, clinical and epidemiological characteristics, study medicine tolerability, adherence, and outcomes will be recorded online through self-report captured in internet-based questionnaires (administered in REDCap). All participant submitted data will be stored and maintained in a secure server. Data from the United States (US) will be stored on the secure servers at the University of Minnesota, and data from Canada will be stored on the secure servers at the Research Institute of the McGill University Health Centre. Both the US and Canadian databases will be stored and managed in their respective countries. For interim data safety monitoring board purposes and also upon completion of recruitment, confidential de-identified data will be shared to facilitate analysis through institutional data sharing agreements. No paper documents will be retained or stored.

Enrollment progress reports will be generated for each trial at 25%, 50%, and 75% of enrollment to highlight the number of participants enrolled, on study, completed, or lost to follow up. The cumulative incidence of COVID-19 (both arms pooled) and cumulative hospitalizations (both arms pooled) will be reported.

### Sample size

According to existing research, the attack rate of transmission to household contacts is approximately 10%.^18^ We used Fisher’s Exact Test and a two-sided α = 0.05 and test power of β = 0.90 to detect a 50% relative risk reduction in disease incidence. Using 1:1 randomization, we calculated a required sample size of n=621 per arm. Accounting for a predicted 20% dropout rate with an internet self-study design the sample size was inflated to n=746 per arm. Therefore, we aimed to recruit a total of 1500 patients for the PEP trial. For symptomatic patients, the proportion of patients who progress to require hospitalization is approximately 10%.^14^ For this trial, we assumed that without treatment 90% of the control arm would not be hospitalized, 8% would be hospitalized without an ICU stay or death, and 2% would be hospitalized with an ICU stay or death. With 1:1 randomization and 1464 total participants, the PET trial has 90% power to detect a log odds ratio of 0.70. A number of sample size approximations were made accounting for differences in baseline rates and relative effect sizes. These can be found in the Appendix 1 and 2.

### Statistical methods

Primary and secondary outcomes will be analyzed separately in PEP and PET trials by intention-to-treat.

Primary outcome for PEP:

The incidence of COVID-19 disease by day 14 will be assessed in the intervention vs. control arm by the Fisher’s Exact Test.

Primary outcome for PET:

The ordinal scale for disease severity (outpatient; hospitalized without ICU; hospitalized with ICU; death) at day 14, will be assessed in the intervention vs. control arms by proportional odds model.

Secondary outcomes (PEP and PET are analyzed separately):

In the PEP group, for the subset of patients who develop symptoms of COVID-19 <u>after</u> taking at least one dose of the study medicine, we will analyze the change in symptom severity in the intervention vs. control group. We will use the same approach as for the primary outcome in the PET group. Participants who are randomized in the PEP group but develop symptoms <u>before</u> taking one dose of the study medicine will be described and analyzed separately in a sub-group analysis.

Secondary outcomes for incidence will be reported with Fisher’s Exact Tests. Continuous valued secondary outcomes will be reported by mean and standard deviation or median with interquartile ranges, depending on their normality. Analysis will be conducted by parametric or non-parametric tests as appropriate. Symptom severity scores will be recorded on days 0, 1, 3, 5, 10 and 14 using a 10-centimeter continuous visual analogue scale, where participants can electronically control a slider indicating symptom severity, with 0 = no symptoms and 10 = severe symptoms. Measurements will be recorded to the nearest 0.1-centimeter. The severity of symptoms at day 5 will be compared first by categorical analysis (symptoms present yes or no) via Fisher’s Exact Chi Square, and subsequently via the independent two-sample *t* test for symptom severity among those who are symptomatic. If data are non-normally distributed, we will analyze data via the Mann-Whitney U test.

Preplanned Subgroup analyses
1. Participants contact had SARS-CoV-2 confirmed by positive molecular test;
2. Participant had SARS-CoV-2 confirmed by positive molecular test;
3. Exposure in healthcare worker versus household contact;
4. Number of days from the exposure;
5. Decile of age;
6. Sex as a biological variable;
7. Censored subjects in the PEP trial who become symptomatic before taking one dose of the study medicine.

### Data monitoring

A Data Safety Monitoring Board (DSMB) will be formed independent of the sponsor and without competing interests. For each trial (PEP and PET), an interim analysis will be presented to the DSMB once the 14 day follow up is achieved for 25%, 50%, and 75% of trial enrollment. All interim analyses will pool US and Canadian deidentified data to arrive at a generalizable conclusion as early as possible. Only the DSMB and unblinded statisticians will be provided with unblinded interim analysis results. The investigators will have access to only pooled results. A Lan-DeMets spending function analog of the O’Brien-Fleming boundaries for the primary outcome for each group will be provided to the DSMB. The stopping boundaries will be truncated at α = 0.001 (|Z| > 3.09). At each analysis, data will be reviewed for safety/efficacy and for achieving stopping rules. Should stopping rules for the primary outcome be met, the DSMB will review secondary outcomes for consistency such that a clear answer is achieved. In the event of early termination for efficacy, the trial will immediately convert to an open-label observational cohort study.

At the second DSMB review, a sample size re-estimation will occur based on the disease transmission rate in the control group. The *a priori* assumption of 10% transmission risk in close contacts is based on limited data in other jurisdictions. A new sample size estimation will take into account the updated transmission rate in this trial and powered to detect a 50% relative reduction in the primary outcome. Starting with the second DSMB review, the DSMB will be given the conditional power under both the trial design parameters and under the current data. If the conditional power is less than 20%, trial discontinuation may be considered.

### Potential harms

Short-term use of hydroxychloroquine is well-tolerated with a safe track record of use for approved indications since 1955. The most commonly reported side effects include: gastrointestinal upset, nausea, vomiting, diarrhea, headache, skin rash or itching.^27^ Gastrointestinal side effects are minimized when taken with meals and when dosages are separated throughout the day. Participants will be instructed to use these optional strategies to increase tolerance.

While a recent study showed a risk of harm with very high doses of chloroquine,^24^ hydroxychloroquine is much less toxic.^26^ Doses used in this trial (3.8 grams total) are similar to malaria treatment doses (2 grams total), and significantly less than treatment for Coxiella infection or other autoimmune conditions (400-600 mg daily, indefinitely), all demonstrating a long history of safety.^27^ A recent trial in COVID-19 using significantly higher doses (1.2 grams daily × 3 days, then 800 mg daily for 2-3 weeks) demonstrated no significant differences in serious adverse advents, though gastrointestinal side effects were higher at this dose.^28^

The safety of taking hydroxychloroquine will be enhanced by excluding those with pre-existing retinopathy, allergic reactions, cardiac conditions, and certain other medications. All adverse reactions will be documented via self-report during virtual visits. Participants will be provided with contact information to be guided on how to manage adverse effects. There are no predefined plans for post-trial care above the standard of care offered at each center. Participants will be provided with updated public health information on the COVID-19 pandemic and instructions for how to proceed in the event of a medical emergency.

### Auditing

Regulatory agencies, institutional sponsors and REBs are authorized to conduct trial audits throughout.

### Confidentiality and access to data

Interactions with trial participants will be through internet-based REDCap electronic case report forms, conforming to required privacy and server security standards. No participant identifying information will be disclosed in any publication or in any other activities arising from this trial. Anonymized data will be pooled with international collaborators, subject to inter-institutional agreements. No information concerning this trial will be released to unauthorized third parties without the prior written approval of the participants, except for monitoring by the REBs or public health authorities. Only immediate study personnel will be authorized to access the database.

### Dissemination policy

Any protocol amendments will be reviewed with each REB and updated on the trial registry and websites. Trial enrollment and results, when available, will be updated on www.clinicaltrials.gov and will be publicly available on the trial websites (www.covidpep.umn.edu and www.covid-19research.ca). Due to the nature of this pandemic, pooled and anonymized trial results will be published immediately, once available, on open access websites and will be submitted urgently for accelerated peer reviewed publication. Full de-identified data will be made available to qualified researchers upon request.

## Discussion

Hydroxychloroquine has garnered unprecedented attention as a potential therapeutic option in the global COVID-19 pandemic.^39^ Several *in vitro* and *in vivo* studies have shown data that may support its possible efficacy, but clinical data is limited to small clinical trials and uncontrolled case series and cohorts.^20^ There is an urgent need for the rigorous evaluation of hydroxychloroquine as a therapeutic option against COVID-19.^40^

This innovative trial is adequately powered to rapidly answer urgent questions regarding the effectiveness of hydroxychloroquine and its ability to prevent or reduce the severity of COVID-19. The novel web-based design of this trial, in the setting of a highly contagious global pandemic, has ensured that all interactions with participants, including enrollment and informed consent, are done remotely using courier services for medicine delivery and internet-based follow-up. This will allow compliance with strict social distancing strategies and reduce risks of infectious spread to study personnel and others. The design encourages large scale enrollment for rapid achievement of enrollment target and trial results. Importantly, the design was finalized, regulatory and REB approvals were obtained, and large geographic jurisdictions brought online in an unprecedently short duration (within a few weeks of a global pandemic) with a goal to obtain the maximal information in the minimal amount of time.

Recently, several public figures have promoted the widespread use of hydroxychloroquine in the absence of robust scientific data.^39,41^ This trial will help address if large scale dissemination of hydroxychloroquine is appropriate in certain populations. Importantly, if this study shows a positive effect, this will support a new therapeutic option to mitigate the global spread and impact of COVID-19. If a null or negative effect is shown, this will reduce the risk of harm to future patients, reduce health care expenditures on ineffective medications, and protect the drug supply for patients with previously established indications for hydroxychloroquine. The results of this trial will be instrumental in determining if hydroxychloroquine may play an important role in mitigating the global impact of COVID-19.

## Data Availability

Data will be provided when the full trial results are published

---

**Appendix 1:** Sample size calculation table for the post-exposure prophylaxis study using the Fisher’s Exact test

| **Estimated transmission rate with placebo** | **Estimated transmission rate with drug** | **Percent relative reduction in transmission rate** | **Sample size (per arm) with 90% power** | **Sample size (per arm) with 80% power** |
| --- | --- | --- | --- | --- |
| 10% | 5% | 50% | 621 | 474 |
|  | 6% | 40% | 1014 | 771 |
| 12% | 6% | 50% | 509 | 389 |
|  | 7.2% | 40% | 831 | 632 |
| 15% | 7.5% | 50% | 398 | 304 |
|  | 9% | 40% | 648 | 493 |

**Appendix 2:**
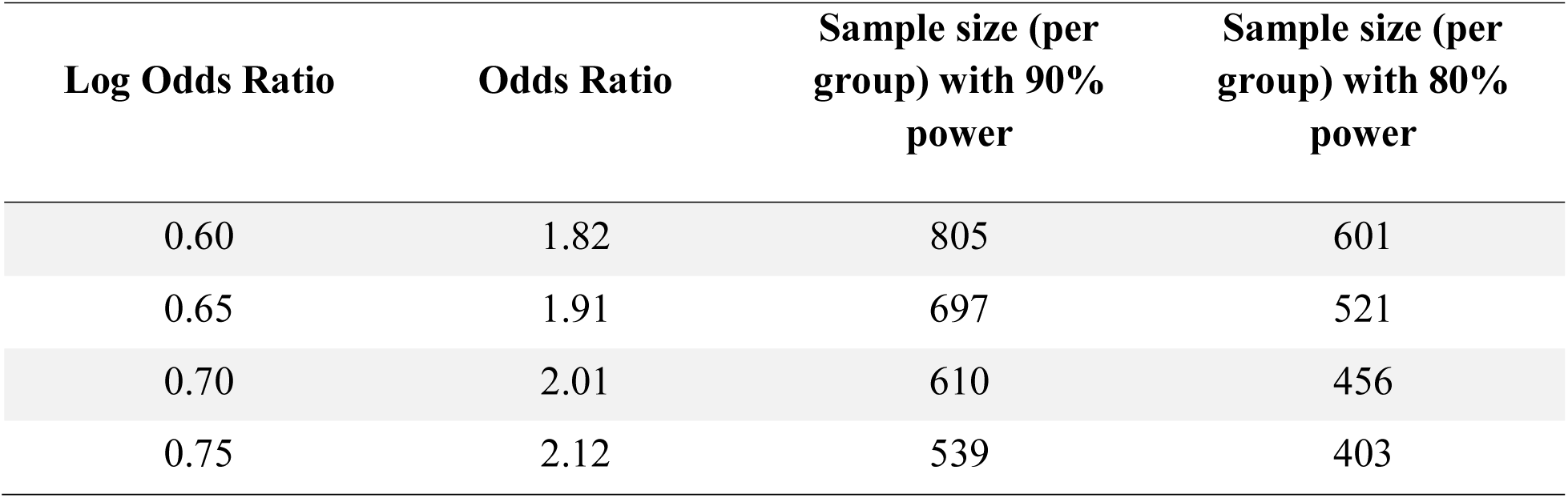
Sample size calculation table for the pre-emptive therapy study using the Fisher’s Exact test

